# The impact of social distancing on COVID19 spread: State of Georgia case study

**DOI:** 10.1101/2020.04.29.20084764

**Authors:** Pinar Keskinocak, Buse Eylul Oruc, Arden Baxter, John Asplund, Nicoleta Serban

## Abstract

As the spread of COVID19 in the US continues to grow, local and state officials face difficult decisions about when and how to transition to a “new normal.” The goal of this study is to project the number of COVID19 infections and resulting severe outcomes, and the need for hospital capacity under social distancing, particularly, shelter-in-place and voluntary quarantine for the State of Georgia. We developed an agent-based simulation model to project the infection spread. The model utilizes COVID19-specific parameters and data from Georgia on population interactions and demographics. The simulation study covered a seven and a half-month period, testing different social distancing scenarios, including baselines (no-intervention or school closure only) and combinations of shelter-in-place and voluntary quarantine with different timelines and compliance levels. The following outcomes are compared at the state and community levels: the number and percentage of cumulative and daily new symptomatic and asymptomatic infections, hospitalizations, and deaths; COVID19-related demand for hospital beds, ICU beds, and ventilators. The results suggest that shelter-in-place followed by voluntary quarantine reduced peak infections from approximately 180K under no intervention and 113K under school closure, respectively, to below 53K, and delayed the peak from April to July or later. Increasing shelter-in-place duration from four to five weeks yielded 2-9% and 3-11% decrease in cumulative infection and deaths, respectively. Regardless of the shelter-in-place duration, increasing voluntary quarantine compliance decreased daily new infections from almost 53K to 25K, and decreased cumulative infections by about 50%. The cumulative number of deaths ranged from 6,660 to 19,430 under different scenarios. Peak infection date varied across scenarios and counties; on average, increasing shelter-in-place duration delayed the peak day by 6 days. Overall, shelter-in-place followed by voluntary quarantine substantially reduced COVID19 infections, healthcare resource needs, and severe outcomes.

## Introduction

The novel coronavirus SARS-CoV-2 causes a rapidly spreading respiratory illness, Coronavirus Disease 2019 (COVID19), which has become a pandemic [1]. During the early stages of a pandemic, medical interventions, such as vaccines or antiviral treatments, are either non-existent or extremely limited [2]. Hence, local, national, and global governments and public officials wrestle with the difficult decisions of how, when, and where to implement non-medical interventions [3]. The decision-makers also need to understand how the type and duration of interventions, as well as the public’s compliance levels, impact their effectiveness [4].

In this study, we developed an agent-based simulation model to predict the spread of COVID19 geographically and over time. The model captures both the natural history of the disease and interactions in households, workplaces, schools, and communities [5-9]. The model was populated with COVID19 parameters from the literature and population-related data from the State of Georgia, including demographic information, household sizes, and travel patterns, and validated using data regarding COVID19 confirmed infections and deaths in Georgia. The model’s outputs include new daily infections (symptomatic and asymptomatic by age group), hospitalizations, and deaths at the census tract level.

We utilized the model to evaluate the effectiveness and impact of non-medical social-distancing interventions, including school closure, shelter-in-place (SIP), and voluntary quarantine (VQ) [6, 10-16]. We tested various scenarios with different durations and time-varying compliance levels for interventions to inform decision-makers about potential social distancing recommendations to be shared with the public. We also developed a hospital resource estimation decision-support tool, which takes as input the model’s daily COVID19-related hospitalization estimates, and predicts the number of hospital beds, ICU beds, and ventilators needed geographically (at the county level) and over time. We then aggregated these estimates across the fourteen coordinating hospital regions in Georgia, to provide insights about potential capacity shortages in the healthcare system [17].

## Methods

### Study population

Population in Georgia stratified by age groups 0-4, 5-9, 10-19, 20-64, 65+. In Georgia there are 1,336,490, 1,418,910, 6,685,870, and 1,356,730 people in age groups 0-9, 10-19, 20-64, and 65 or over, respectively, with a total population of approximately 10,519,000 [18, 19].

### Infection projection model

We adapted an agent-based simulation model with heterogeneous population mixing to predict the spread of the disease geographically during the study period of February 18, 2020 to September 30, 2020 [5-8]. The model captures the natural history of the disease at the individual level, by age group, as well as the infection spread via a contact network consisting of interactions in households, peer groups (workplaces, schools), and communities, with different rates of transmission [13, 20-25].

The model was populated with COVID19-specific parameters [20-22, 26-40] and data from Georgia, including household type [18], household size [18], children status [18, 41], workflow [42], and population demographics [18] at the census tract level. To seed the model, we utilized the confirmed infection data for Georgia, at the county level [43].

The main assumptions in the model were (1) every individual is in one of the following states at any given time (see Fig 1): susceptible (S), exposed (E), transition (IP), asymptomatic (IA), symptomatic (IS), hospitalized (H), recovered (R), or dead (D) and (2) three levels of mixing in the population: (i) community (day and night), (ii) peer groups (day), and (iii) household (night). During the exposed state, an infected individual shows no symptoms and does not infect others. During the transition state an infected individual shows no symptoms but could infect others. From the transition state, an infected individual moves to the symptomatic or the asymptomatic state.

**Fig 1.**
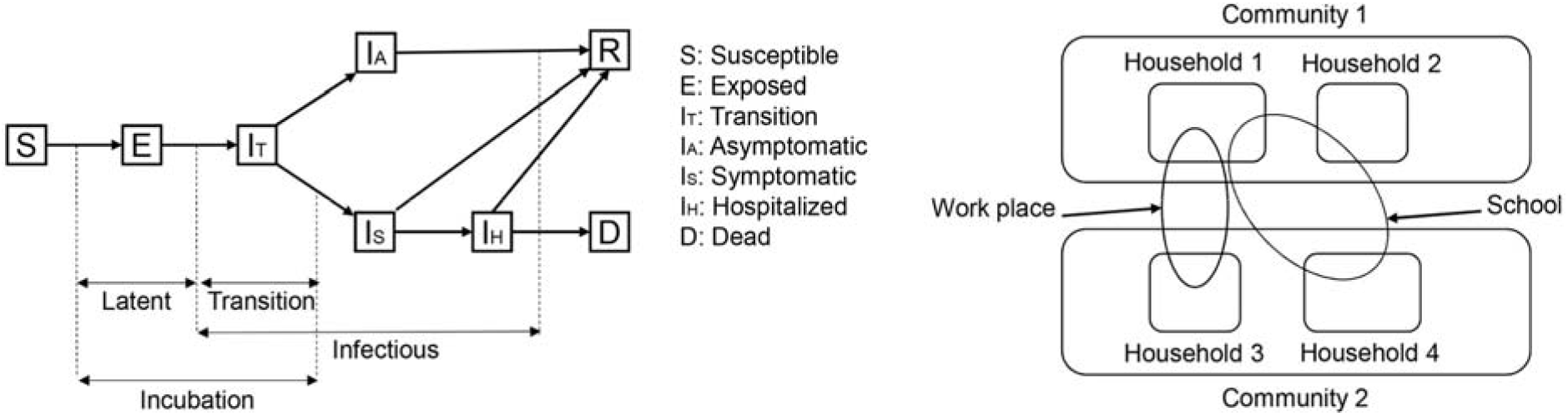
Model descriptions. Agent-based model incorporates the natural history of the disease for each individual agent, by age group, and the interactions at the household, peer group, and community, across different geographic areas. Outcome measures reported are averages of 30 replications ran for each scenario.

S1 Appendix provides additional details on the model implementation. Table 1 provides the input model parameters, and Fig A in S1 Appendix provides model validation using COVID19 confirmed cases and deaths reported in Georgia.

**Table 1.**
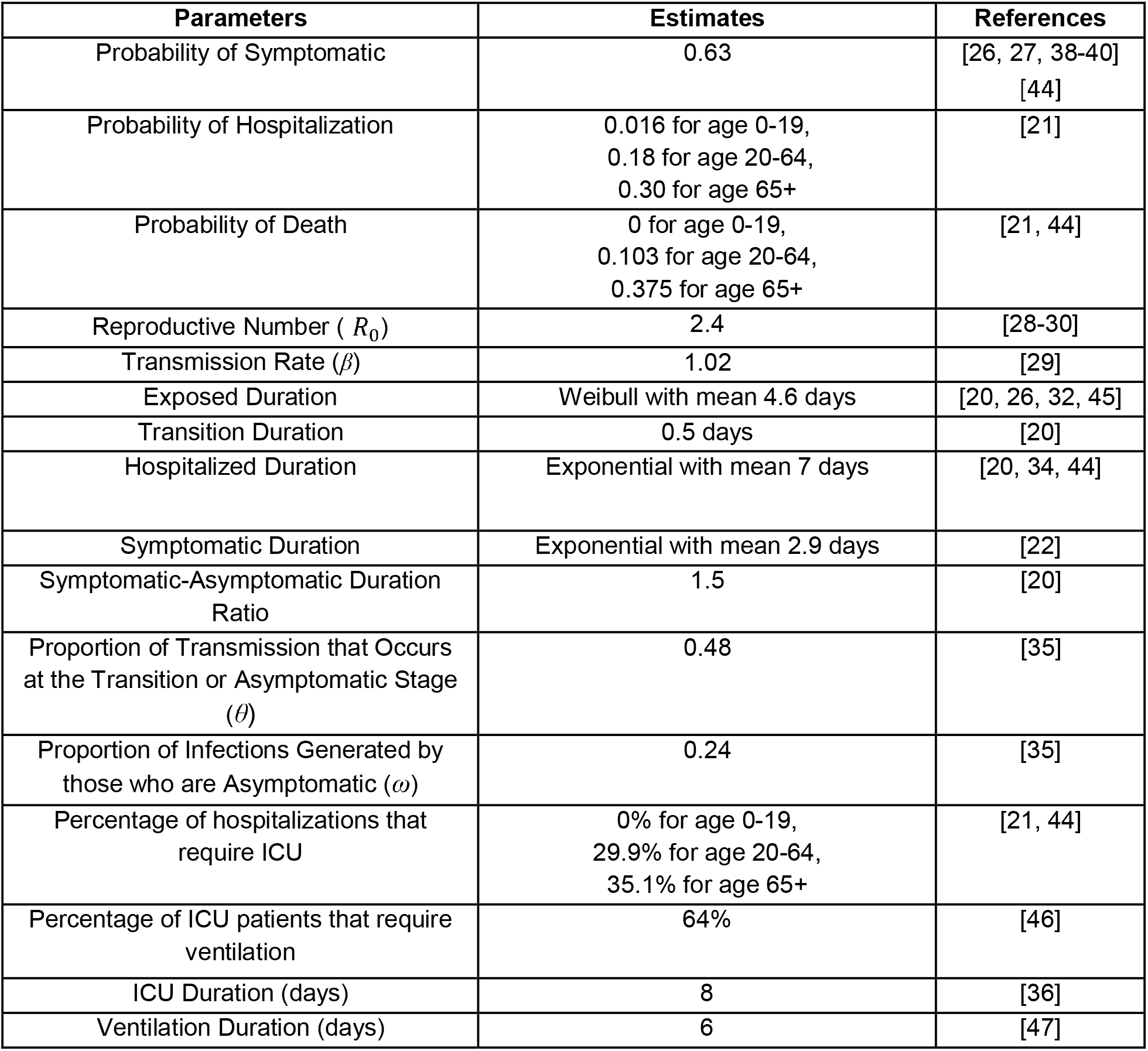
Model parameters. Descriptions and references for the model input parameters.

| Parameters | Estimates | References |
| --- | --- | --- |
| Probability of Symptomatic | 0.63 | [26, 27, 38-40]<br>[44] |
| Probability of Hospitalization | 0.016 for age 0-19,<br>0.18 for age 20-64,<br>0.30 for age 65+ | [21] |
| Probability of Death | 0 for age 0-19,<br>0.103 for age 20-64,<br>0.375 for age 65+ | [21, 44] |
| Reproductive Number ( $R_0$ ) | 2.4 | [28-30] |
| Transmission Rate ( $\beta$ ) | 1.02 | [29] |
| Exposed Duration | Weibull with mean 4.6 days | [20, 26, 32, 45] |
| Transition Duration | 0.5 days | [20] |
| Hospitalized Duration | Exponential with mean 7 days | [20, 34, 44] |
| Symptomatic Duration | Exponential with mean 2.9 days | [22] |
| Symptomatic-Asymptomatic Duration Ratio | 1.5 | [20] |
| Proportion of Transmission that Occurs at the Transition or Asymptomatic Stage ( $\theta$ ) | 0.48 | [35] |
| Proportion of Infections Generated by those who are Asymptomatic ( $\omega$ ) | 0.24 | [35] |
| Percentage of hospitalizations that require ICU | 0% for age 0-19,<br>29.9% for age 20-64,<br>35.1% for age 65+ | [21, 44] |
| Percentage of ICU patients that require ventilation | 64% | [46] |
| ICU Duration (days) | 8 | [36] |
| Ventilation Duration (days) | 6 | [47] |

### Intervention analysis

The following baseline scenarios and social distancing interventions are analyzed in our study:

1. *<u>No intervention (NI)</u>* – the population interacts with each other normally;
2. *<u>School Closure (SC)</u>* – no peer group interactions among children (i.e., no K-12 school interactions);
3. *<u>Voluntary Quarantine (VQ)</u>* – All household members stay home if a household member experiences symptoms, until the entire household is symptom-free.
4. *<u>Shelter-in-Place (SIP)</u>* – Household members stay home complying with a state order.
5. *<u>Voluntary Shelter-in-Place (VSIP)</u>* – Household members choose to follow SIP voluntarily.

Household members complying with SIP, VSIP, or VQ do not engage in peer group or community interactions. Compliance levels (<100%) under SIP, VSIP, and VQ probabilistically determine individual compliance and corresponding community interactions.

NI and SC were considered as *baselines* for comparison. In Scenarios 1-9, SIP durations (4, 5, and 6 weeks) and gradually decreasing post-SIP VQ compliance levels (low, medium, high) were tested (Fig 2*); s*helter-in-place was in effect for 4 weeks (April 3-April 30) in Georgia [48]. Because all K-12 schools in Georgia were closed starting March 16, 2020 until the end of July, Scenarios 1-9 assumed school closure.

**Fig 2.**
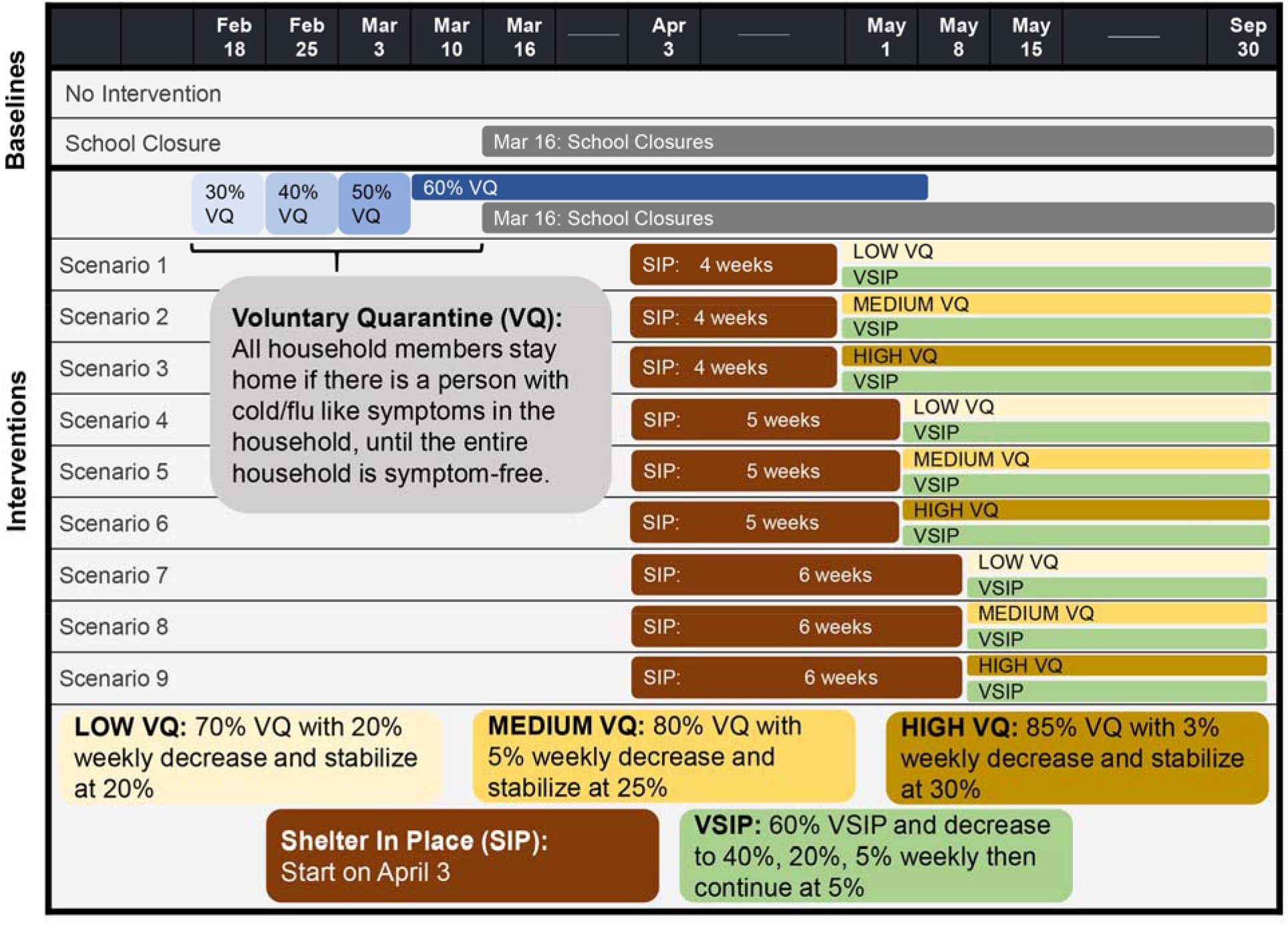
Intervention scenarios. Description of the intervention scenarios considered in this study.

Additionally, after the end of SIP in Scenarios 1-9, decreasing compliance with voluntary shelter-in-place was considered, chosen to be in line with social mobility indicators [49]. Further details on the choice of VSIP compliance levels can be found in S1 Appendix.

### Healthcare resource needs projection model

The hospitalization output from the simulation model was used to estimate the daily demand for hospital beds (general inpatient beds and intensive care unit (ICU) beds) and ventilators for COVID19 patients. Daily hospital bed demand was calculated by aggregating the number of hospital beds needed in the previous day with the number of new hospitalizations, minus the proportion of the population that was discharged from the hospital based on the average hospitalization duration. ICU bed and ventilator demand were estimated using the percentage of hospitalized patients that require ICU (by age), average ICU duration, percentage of ICU patients that require ventilation, and the average ventilation duration.

We derived county-level risk factors (see Fig B in S1 Appendix) by applying the principal component analysis [50] on several factors known to impact a higher risk of complications and severe outcomes for COVID19 infections, including prevalence of asthma, diabetes, obesity, smoking, cardiovascular disease and chronic conditions [51]. We then adjusted the estimated demand using these risk factors.

S1 Appendix and Table 1 provide additional details on the estimation approach and the input model parameters along with their references.

### Outcome measures

The outcome measures considered for the study period include:

- *New Infection Count (NIC):* number of daily new symptomatic and asymptomatic infections.
- *Infection attack rate (IAR):* cumulative percentage of the population infected.
- *Peak infection (PI):* maximum percentage of the population infected on a given day.
- *Peak day*: The day when NIC is highest.
- *Clinical attack rate (CAR):* percentage of symptomatic infections among the total population.
- *Hospital Bed Demand (HB):* number of hospital beds (general inpatient and ICU beds) needed due to severe outcomes among the infected.
- *Intensive Care Unit Bed Demand (ICUB):* number of ICU beds needed due to severe outcomes among the infected.
- *Ventilator Demand (V):* number of ventilators needed due to severe outcomes among the infected.

## Results

### State-level outcome measure analysis

Fig 3 shows the NIC outcome for all scenarios. Table 2 includes summaries across all scenarios and outcomes. Fig A in S1 Appendix and S1, S2 Figs provide state-level outcomes for Scenarios 1-9.

**Fig 3.**
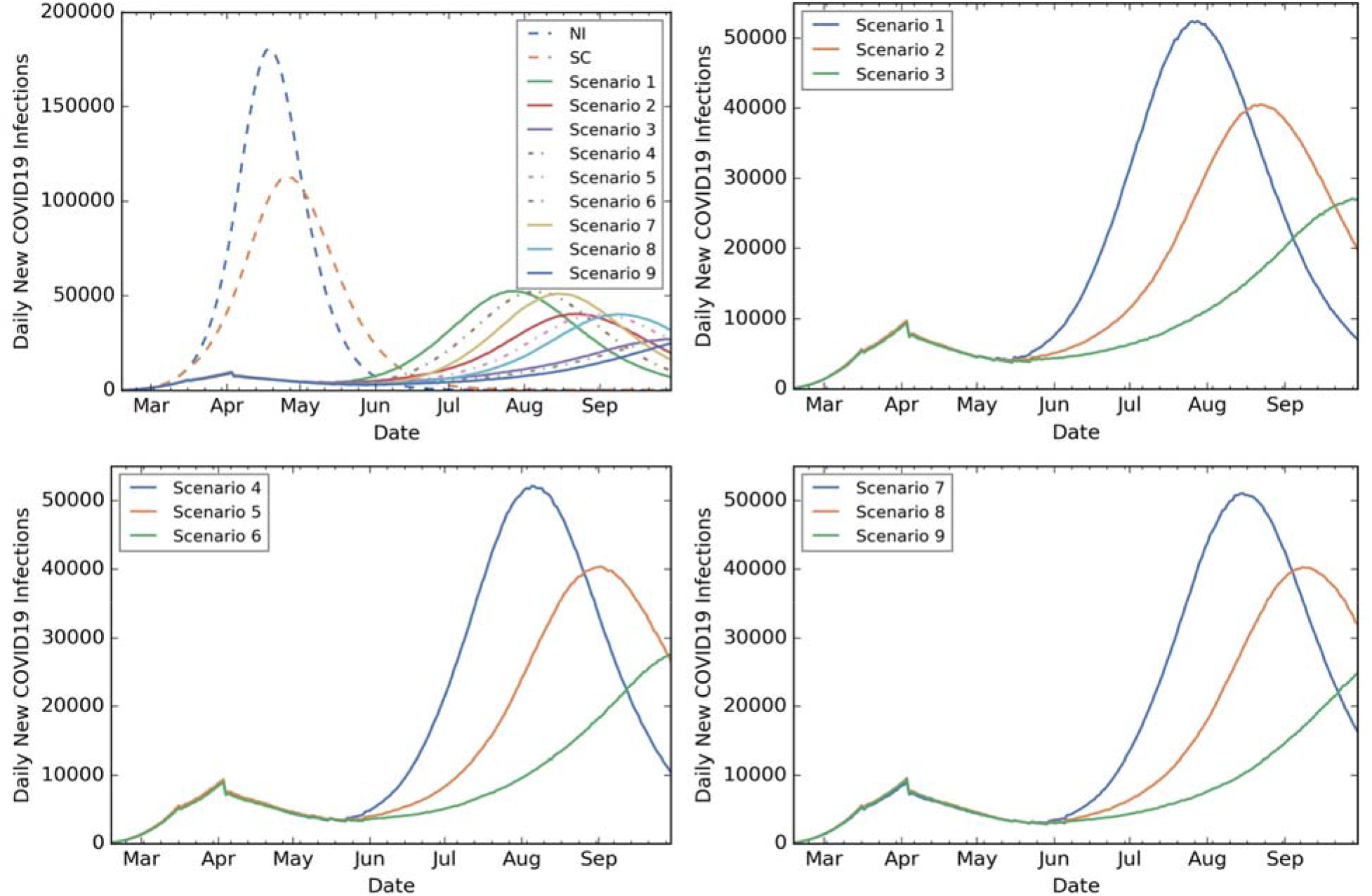
State level outcomes: NIC under all scenarios; 4-week, 5-week and 6-week SIP scenarios. NIC under all scenarios including baseline (top left plot), NIC under 4week SIP followed by Low (Scenario 1), Medium (Scenario 2), High (Scenario 3) VQ compliance (top right plot), NIC under 5-week SIP followed by Low (Scenario 4), Medium (Scenario 5), High (Scenario 6) VQ compliance (bottom left plot), NIC under 6week SIP followed by Low (Scenario 7), Medium (Scenario 8), High (Scenario 9) VQ compliance (bottom right plot).

**Table 2.**
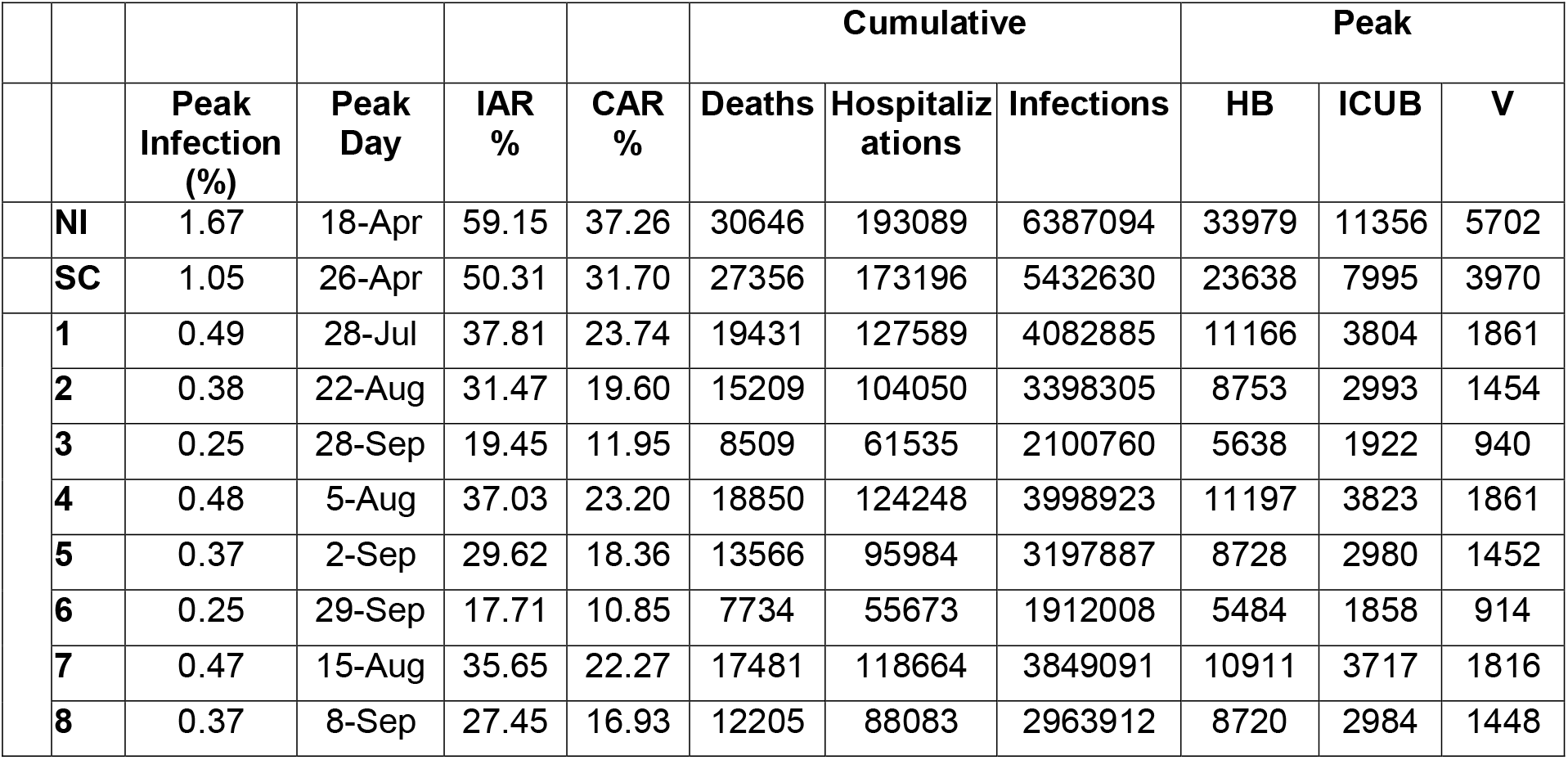
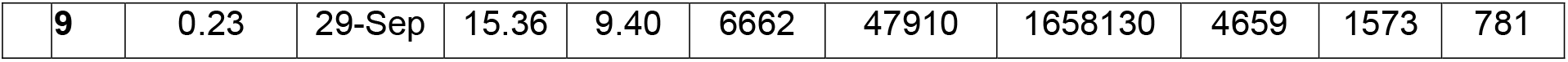
Statewide outcome measures. Statistical summaries that compare baseline and intervention scenarios with respect to Peak Infection (%), Peak Day, IAR (%), CAR (%), Cumulative Deaths, Cumulative Hospitalization, Cumulative Infections, Peak HB, Peak ICUB, and Peak V.

The maximum NIC was approximately 180K (April 18th) and 113K (April 26th) under NI and SC, respectively. Under Scenarios 1-9, the maximum NIC was below 53K, with the earliest peak in July. Compared to NI and SC, in Scenarios 1-9 NIC was at least 36% and 25% lower, and approximately 2.3 million and 1.4 million fewer people infected, respectively. Extending SIP by 1-2 weeks or following SIP by high VQ further reduced NIC and delayed the peak day. Similar trends were observed for CAR, with the number of symptomatic infections reducing by more than 36% and 25%, compared to NI and SC, respectively.

Increasing the SIP duration from four to five weeks (i.e., extending by one week) yielded a decrease of approximately 2-9% in IAR and CAR, approximately 3-10% in cumulative hospitalizations, and 3-11% in cumulative deaths. Increasing the SIP duration from five to six weeks (i.e., extending by two weeks) yielded a decrease of approximately 4-13% in IAR, 4-14% in cumulative hospitalizations, and 7-14% in cumulative deaths.

Higher VQ compliance after SIP, regardless of the SIP duration, decreased the peak NIC from approximately 53K to 25K and decreased IAR by up to 57%.

SC had a lower IAR outcome than NI, but led to similar severe outcomes (cumulative deaths and hospitalizations). Healthcare resource demand estimates (HB, ICUB, and V) were about 30% lower under SC versus NI. In the absence of social distancing interventions, approximately 30,640 and 27,350 people in Georgia were projected to die, and approximately 193,080 and 173,190 people were projected to be hospitalized under NI and SC, respectively.

The total number of deaths for Scenarios 1-9 ranged from 6,662 (Scenario 9) to 19,431 (Scenario 1); the number hospitalized was between 47,910 (Scenario 9) and 127,589 (Scenario 1); peak hospital bed needs ranged from 4,659 (Scenario 9) to 11,166 (Scenario 1). Peak ICU bed and ventilator needs ranged from 1,573 (Scenario 9) to 3,804 (Scenario 1) and from 781 (Scenario 9) to 1,861 (Scenario 1), respectively. Scenario 9, where SIP extended until mid-May followed by high VQ, provided the lowest cumulative infections, hospitalizations, and deaths.

### Infection spread outcome measure analysis by county

Tables A and B in S1 Appendix include the estimated peak day and percentage by county across all scenarios. S3 Fig includes the maps of the NIC by county for different dates.

Peak day varied across scenarios and counties; however, on average, increasing SIP duration by one week moved the peak day by 6 days across counties. Each week of SIP extension delayed the peak day by an average of 6-7 days in the most populated counties (e.g., Fulton, Gwinnett, Cobb, DeKalb, Chatham) and 3-6 days in smaller counties (e.g., Glascock, Clay, Webster, Quitman, Taliaferro), respectively. Increasing VQ compliance from low to medium and from medium to high delayed the peak day by an average of 24 and 19 days, respectively. In most scenarios, rural counties peaked on average 5 days later compared to urban counties. Differences in the peak day observed in rural and urban counties were mostly consistent across the scenarios.

PI fluctuated depending on SIP duration. Rural counties and urban counties did not differ much in terms of PI. See S1 Appendix for examples of an analysis of urban and rural counties in Georgia.

The NIC was highest in the densely populated Fulton county and other surrounding counties in the Atlanta metropolitan area across all scenarios.

### Healthcare resource needs analysis by coordinating hospital region

Fig 4 presents the healthcare resource peak demand under Scenario 2, by hospital region. (A map of the 14 coordination hospital regions of Georgia can be found in [52].) Fig 5 shows the hospital and ICU bed needs over time for region N under Scenario 2. Similar patterns were observed for other scenarios.

**Fig 4.**
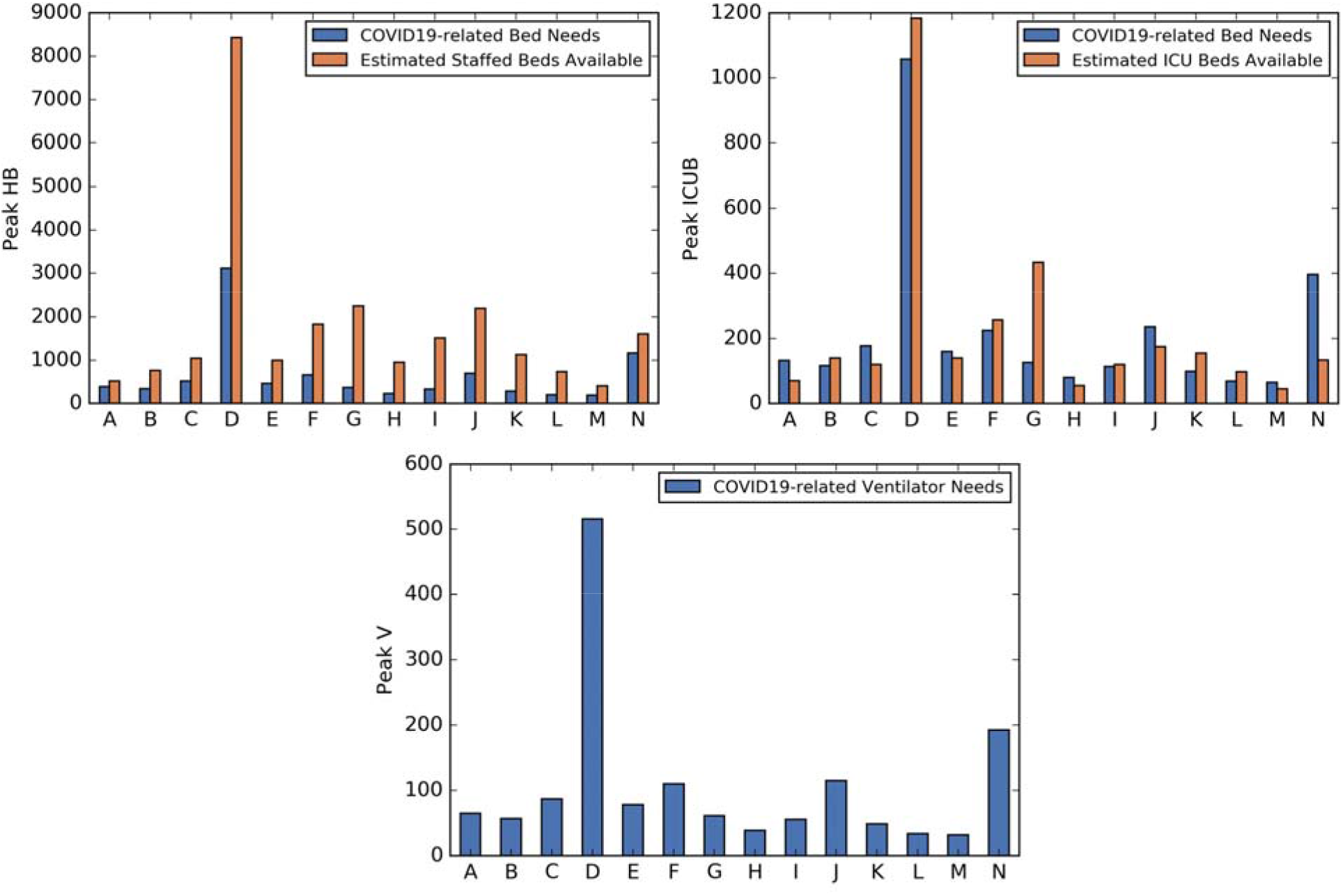
COVID19-related peak healthcare resource demand for 4-week SIP, Medium VQ compliance post-SIP. COVID19-related hospital bed (top left plot), ICU bed (top right plot), and ventilator (bottom center plot) peak needs for 4-week SIP, Medium VQ compliance post-SIP (Scenario 2).

**Fig 5.**
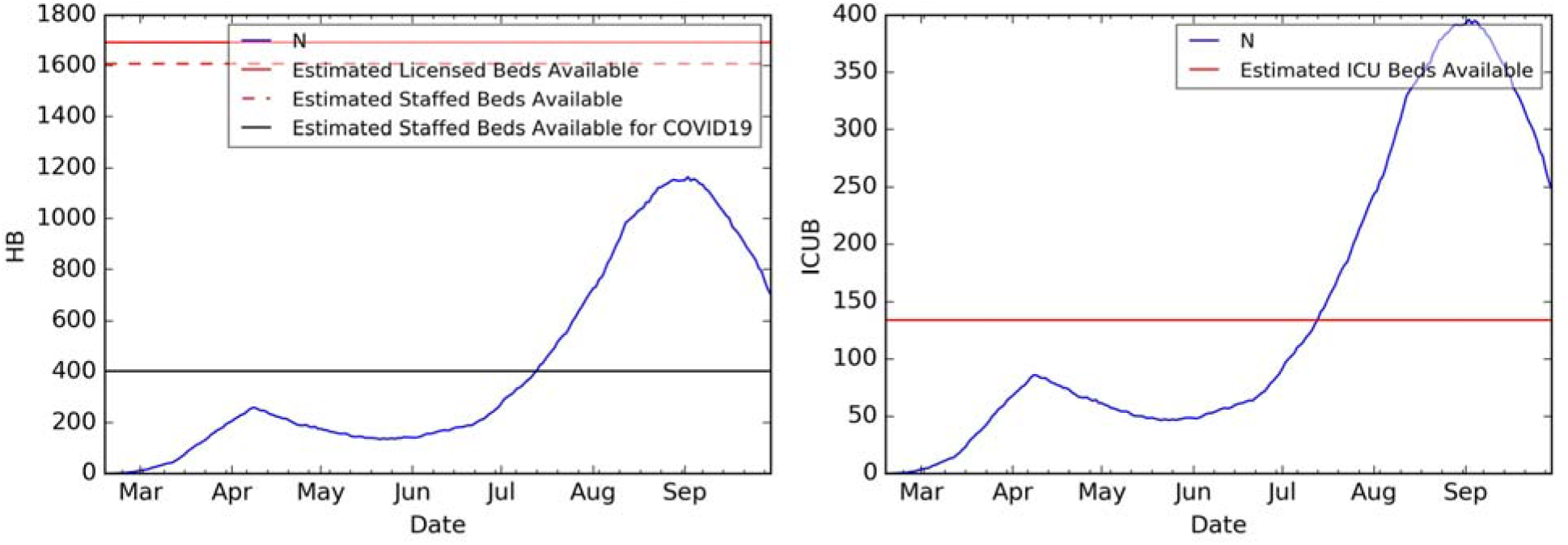
COVID19-related healthcare resource demand over time for region N for 4week SIP, Medium VQ compliance post-SIP. COVID19-related hospital bed (left plot) and ICU bed (right plot) needs over time for region N under 4-week SIP, Medium VQ compliance post-SIP (Scenario 2).

The highest need for COVID19-related healthcare resources was in region D, with 3,117 hospital beds, 1,058 ICU beds, and 516 ventilators. Region D includes three of the top four populous counties in Georgia. The four regions with the highest need across all outcomes were regions D, F, J, and N, which are regions that include populous counties.

There is a gap between HB and ICUB demand and availability around the peak day in many regions. For example, consider region N under Scenario 2. The peak hospital bed need for COVID19 patients is projected to occur around September 2^nd^ with a demand of 1,164 beds, yet the estimated staffed hospital bed availability for all (including nonCOVID19) patients is 1,607 [53]. Peak ICU bed demand for COVID19 patients was projected as 396, also occurring around September 2^nd^ whereas estimated availability for all patients is 134.

## Discussion

During the early phases of a pandemic, in the absence of a vaccine or effective treatments, non-medical interventions are of utmost importance. During COVID19, the majority of schools in the US closed around mid-March for the remainder of the school year [54], and governors issued shelter-in-place orders [55] during March or April. Many states ended their shelter-in-place orders towards the end of April or early May, given the financial, social, and psychological impacts of shelter-in-place. For example, in Georgia, schools closed on March 16, 2020 [56], shelter-in-place orders were issued on April 3, 2020, transitioning out of shelter-in-place started on April 24, 2020 [57], and shelter-in-place officially ended on April 30, 2020. The number of new COVID19 confirmed infections in Georgia have rapidly increased since early June [46]. In this paper, we analyzed the impact of shelter-in-place duration and social distancing compliance levels, particularly voluntary quarantine, using data from Georgia.

For baseline comparisons, we ran two scenarios: no intervention and school closure only. We tested nine intervention scenarios, assuming school closures starting on March 16, 2020, followed by shelter-in-place on April 3, 2020. In these nine scenarios, we modeled a slowly increasing social distancing compliance prior to school closures, shelter-in-place durations of 4-6 weeks, voluntary shelter-in-place and voluntary quarantine compliance levels of low, medium, and high after the end of shelter-in-place. Our study period extended through the end of September.

Compared to Scenarios 1-9, infections are higher in the baseline scenarios, with peak infections occurring around mid-to-late April. Scenarios 1-9 show that shelter-in-place could significantly slow down the disease spread, protecting public health, and offering the opportunity for better preparedness of healthcare resource capacity. Each week extension of shelter-in-place (beyond 4 weeks) could delay the peak day by about 6 days.

Social distancing (modeled by voluntary quarantine compliance) showed a significant impact on all outcome metrics, particularly, peak day and peak infections. Depending on social distancing compliance levels, the peak infections under low compliance levels could exceed 50K. State level peak percentage could also decrease significantly as the voluntary quarantine compliance increases, observing the same effect of post shelter-inplace compliance at the county level as well.

After the end of shelter-in-place, voluntary quarantine compliance had a significant impact on COVID19-related deaths, which could approach 20K by the end of September in the case of low voluntary quarantine compliance. The death numbers could be even higher because the demand would significantly exceed healthcare capacity, especially around the peak.

Infection spread across counties varied over time, with earlier peak days in some counties compared to others, which impacts resource allocation decisions across the state. For example, in Scenario 1 (shelter-in-place 4 weeks, low voluntary quarantine compliance after shelter-in-place), several of the larger counties could reach their peak around late July, with the peak days of other counties extending to late July or mid-August. Similarly, there was a variation among counties in terms of the peak infection percentage under different voluntary quarantine compliance levels.

Estimates for COVID19-related needs for hospital beds, ICU beds, and ventilators suggested shortages in all scenarios across the 14 coordinating hospital regions in Georgia. Even if all available hospital resources were used for COVID19 patients, at the peak, these resources would still not be sufficient for some of the regions. In some regions, the shortage would continue for several weeks. These results further emphasize the importance of voluntary shelter-in-place and high compliance levels for social distancing after the end of state-mandated shelter-in-place.

During shelter-in-place, the withdrawal of a large percentage of the population (e.g., 80%, depending on compliance level) from interactions raises social and economic concerns. Hence, voluntary quarantine is an effective intervention if widely adopted, but significantly less disruptive than shelter-in-place.

Another important advantage of voluntary quarantine is the ease of communication and implementation. When COVID19 diagnostic testing capacity is limited, voluntary quarantine can be implemented based on symptoms. Household members are advised to stay home if there is a person with cold- or flu-like symptoms in the household (even in the absence of testing or confirmation of COVID19), until the entire household is symptom-free. High compliance with voluntary quarantine would reduce not only the spread of COVID19 but would have the side benefit of also reducing the spread of the cold or flu.

## Limitations

Most of the limitations of this study lie in the limited data available regarding COVID19 infection and transmission, and the related parameters (which were drawn from the literature), which impact the natural history and severe outcomes for the study period. While we incorporated Georgia data on household types and sizes, children status, workflow, and population demographics, our model made assumptions about the peerto-peer interactions between different age groups, which impact the virus transmission under various intervention strategies. Testing of a wide range of scenarios enabled a better understanding of the impact of social distancing compliance on COVID19 outcomes. This study did not consider the usage of face coverings; the projected infections and deaths would decrease, but the relative effectiveness of interventions would not change with the use of face coverings.

## Conclusions

As states continuously evaluate the benefits versus social and economic costs of non-pharmaceutical interventions such as school closures and shelter-in-place, our results suggest that there needs to be a very strong messaging to the public about social distancing. It is important to re-emphasize that some people might be infected with little or no symptoms and infect others [58]. Voluntary quarantine is one form of social distancing that is easy to communicate; it reduces infection spread (both from symptomatic and asymptomatic individuals), but does not entirely prevent the spread. There may be households with COVID19 infection, and yet no household member might be experiencing symptoms – these households would not be impacted by voluntary quarantine. Therefore, while it is essential to promote voluntary quarantine, strongly encouraging households to continue voluntary shelter-in-place, to the extent possible, or other forms of social distancing would help slow the spread of COVID19. It is also important to enact policies and guidelines for promoting voluntary quarantine at the local and national levels. Without such policies, compliance will be low, and hence, such interventions will become quickly ineffective.

## Data Availability

The manuscript utilizes publicly available data sources and model parameters from the literature. All sources are cited in the manuscript.

https://dph.georgia.gov/covid-19-daily-status-report

http://data.census.gov

http://data5.ctpp.transportation.org

https://github.com/nytimes/covid-19-data

## Acknowledgement

The authors of this paper are thankful to state of Georgia representatives, including Garry McGiboney from Georgia Department of Education, Laura Edison from Georgia Department of Public Health, and Susan Miller and Natalie Lee from Georgia Geospatial Information Office for their support, guidance, or sharing data sources. The authors would like to thank StreetLight Data, Inc. for providing data that motivated our scenario choices. The authors are also thankful to Melody Shellman, Hannah Lin, Ethan Channel, Pravara Harati, April Zhuoting Yu, Gabriel Siewert, and Christopher Stone for supporting various parts of the projects, and to the anonymous PLoS One reviewers and editors whose constructive comments and suggestions helped significantly improve the content and exposition.

## Supporting information

### S1 Appendix. Data sources, model description and model inputs

**S1 Fig.**
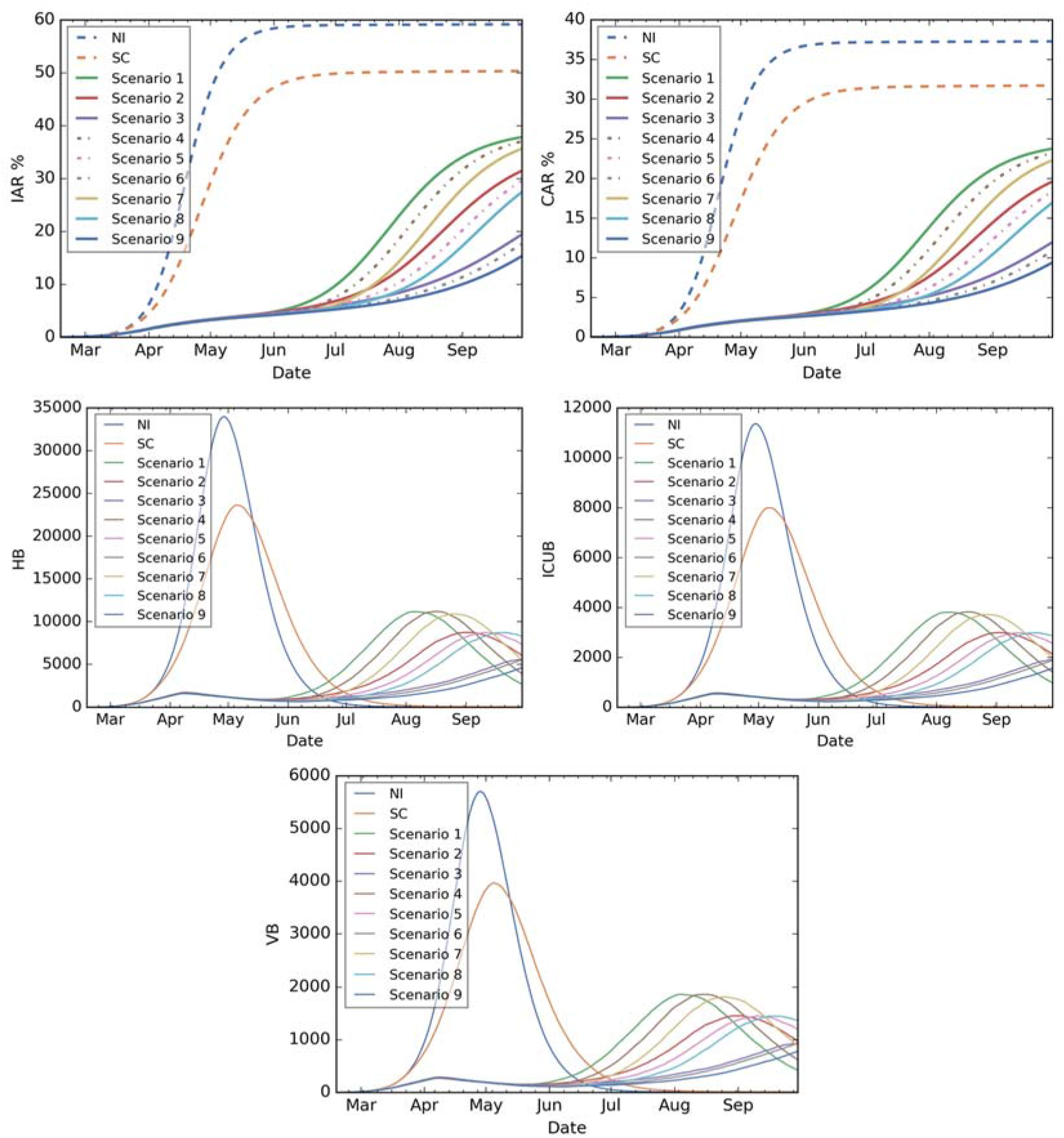
State level outcomes: IAR, CAR, HB, ICUB, V across all scenarios. State Level Outcomes: IAR (first row left plot), CAR (first row right plot), HB (second row left plot), ICUB (second row right plot), V (third row center plot) across all scenarios (including the baseline scenarios).

**S2 Fig.**
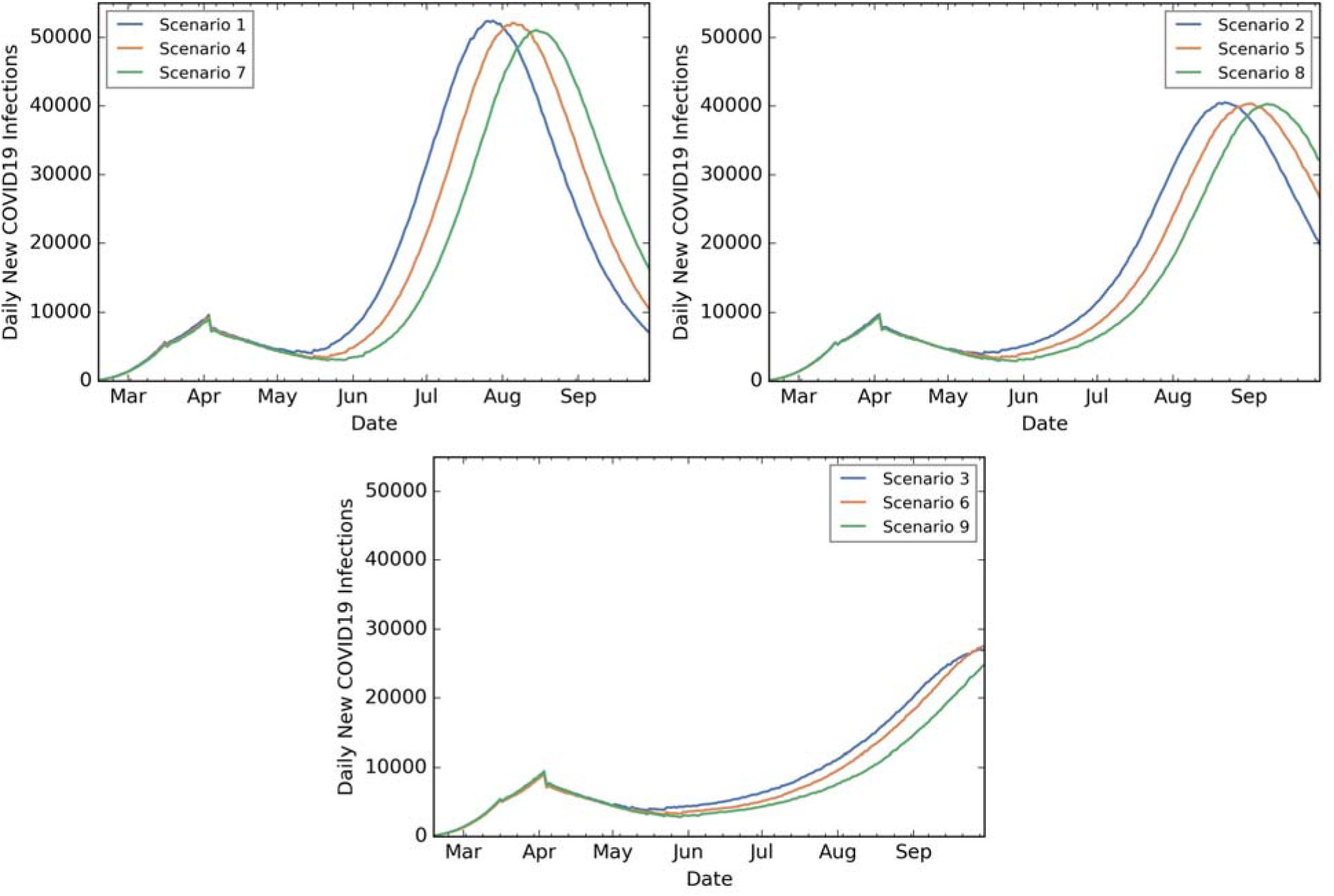
State level outcomes: NIC for low, medium, and high levels of VQ after 4, 5, and 6 weeks of SIP. Daily new COVID19 infections when Low VQ is combined with 4 week (Scenario 1), 5 week (Scenario 4), 6 week (Scenario 7) SIP (top left plot), Medium VQ is combined with 4 week (Scenario 2), 5 week (Scenario 5), 6 week (Scenario 8) SIP (top right plot), High VQ is combined with 4 week (Scenario 3), 5 week (Scenario 6), 6 week (Scenario 9) SIP (bottom center plot).

**S3 Fig.**
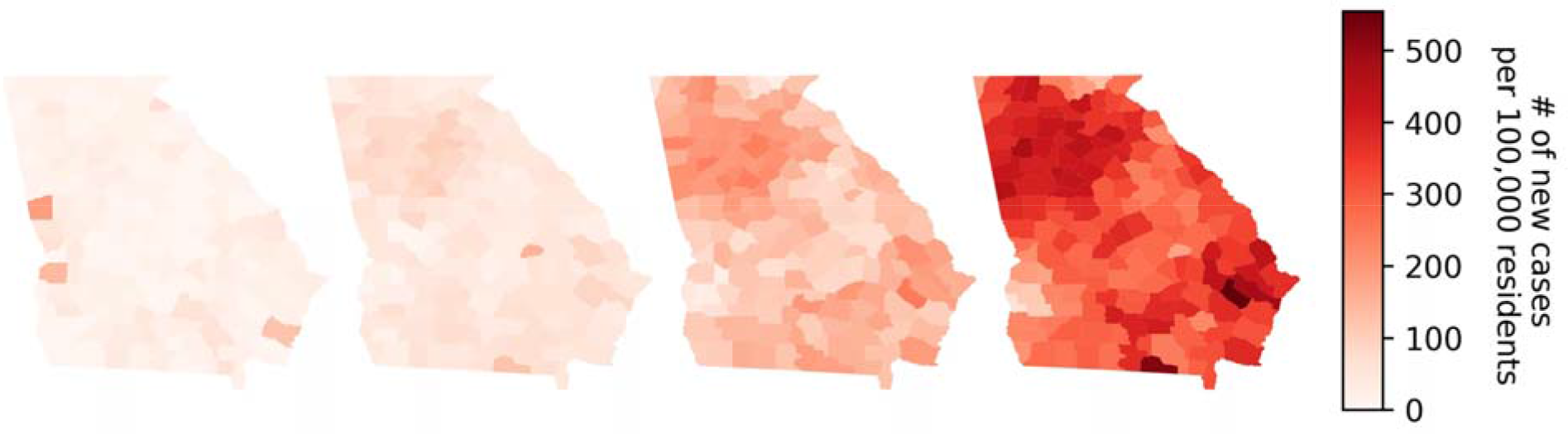
Number of new infections per 100,000 people. Four maps of Georgia at the county level recording the number of new infections per 100,000 people for June 23, 2020 (using the actual number of infections), July 15, 2020 (simulated data from our model), August 15, 2020 (simulated data), and September 15, 2020 (simulated data), respectively [61].

